# The serial interval of COVID-19 from publicly reported confirmed cases

**DOI:** 10.1101/2020.02.19.20025452

**Authors:** Zhanwei Du, Xiaoke Xu, Ye Wu, Lin Wang, Benjamin J. Cowling, Lauren Ancel Meyers

**Affiliations:** The University of Texas at Austin, Austin, Texas 78712, The United States of America; Dalian Minzu University, Dalian 116600, China; Computational Communication Research Center, Beijing Normal University, Zhuhai, 519087, China; School of Journalism and Communication, Beijing Normal University, Beijing, 100875, China; Institut Pasteur, 28 rue du Dr Roux, Paris 75015, France; The University of Hong Kong, Hong Kong SAR, China; Santa Fe Institute, Santa Fe, New Mexico, The United States of America

**Keywords:** Wuhan, coronavirus, epidemiology, serial interval

## Abstract

We estimate the distribution of serial intervals for 468 confirmed cases of COVID-19 reported in 93 Chinese cities by February 8, 2020. The mean and standard deviation are 3.96 (95% CI 3.53-4.39) and 4.75 (95% CI 4.46-5.07) days, respectively, with 12.6% of reports indicating pre-symptomatic transmission.

**One sentence summary:** We estimate the distribution of serial intervals for 468 confirmed cases of COVID-19 reported in 93 Chinese cities by February 8, 2020.

---

A new coronavirus (COVID-19) emerged in Wuhan, China in late 2019 and was declared a public health emergency of international concern by the World Health Organization (WHO) on January 30, 2020 (1). As of February 19, 2020, the WHO has reported over 75,204 COVID-19 infections and over 2,009 COVID-19 deaths (2), while key aspects of the transmission dynamics of COVID-19 remain unclear (3). The serial interval of COVID-19 is defined as the time duration between a primary case (infector) developing symptoms and secondary case (infectee) developing symptoms (4,5). Obtaining robust estimates for the distribution of COVID-19 serial intervals is a critical input for determining the reproduction number which can indicate the extent of interventions required to control an epidemic (6). However, this quantity cannot be inferred from daily case count data alone (7).

To obtain reliable estimates of the serial interval, we obtained data on 468 COVID-19 transmission events reported in mainland China outside of Hubei Province between January 21, 2020, and February 8, 2020. Each report consists of a probable date of symptom onset for both the infector and infectee as well as the probable locations of infection for both cases. The data include only confirmed cases that were compiled from online reports from 18 provincial centers for disease control and prevention (Table S3).

Notably, 59 of the 468 reports indicate that the infectee developed symptoms earlier than the infector. Thus, pre-symptomatic transmission may be occurring, i.e., infected persons may be infectious before their symptoms appear. In light of these negative-valued serial intervals, we find that COVID-19 serial intervals better resemble a normal distribution than more commonly assumed gamma or Weibull distributions (8,9) that are limited to strictly positive values (see Supplement). We estimate a mean serial interval for COVID-19 of 3.96 [95% CI 3.53-4.39] with a standard deviation of 4.75 [95% CI 4.46-5.07], which is considerably lower than reported mean serial intervals of 8.4 days for SARS (9) and 12.6 days (10) - 14.6 days (11) for MERS. The mean serial interval is slightly but not significantly longer when the index case is imported (4.06 days [95% CI 3.55-4.57]) versus locally infected (3.66 days [95% CI 2.84-4.47]); it is slightly shorter when the secondary transmission occurs within a household (4.03 days [95% CI 3.12-4.94]) versus outside of the household (4.56 days [95% CI: 3.85-5.27]). Combining these findings with published estimates for the early exponential growth rate COVID-19 in Wuhan (12,13), we estimate a basic reproduction number (*R*_0_) of 1.32 [95% CI 1.16-1.48] (6), which is lower than published estimates that assume a mean serial interval exceeding seven days (13–15).

These estimates reflect reported symptom onset dates for 752 cases from 93 Chinese cities, who range in age from 1 to 90 years (mean 45.2 years and SD 17.21 years). Recent analysis of COVID-19 case data from mainland China, Taiwan, Hong Kong, Vietnam, South Korea, Germany and Singapore have reported average serial intervals of 7.5 days [95% CI 5.3-19] (13), 4.4 days [95% CI 2.9-6.7] (16) and 4.0 days [95% CrI 3.1-4.9] (17) based on considerably smaller samples of 6, 21 and 28 infector-infected pairs, respectively. Whereas none of these studies report negative serial intervals in which the infectee developed symptoms prior to the infector, 12.6% of the serial intervals in our sample are negative.

We note four potential sources of bias in our estimates, three of which are likely to cause underestimation of COVID-19 serial intervals. First, the data are restricted to online reports of confirmed cases and therefore may be biased towards more severe cases in areas with a high-functioning healthcare and public health infrastructure. The rapid isolation such cases may have prevented longer serial intervals, potentially shifting our estimate downwards compared to serial intervals that might be observed in an uncontrolled epidemic. Second, the distribution of serial intervals varies throughout an epidemic, with the time between successive cases contracting around the epidemic peak (18). To provide intuition, a susceptible person is likely to become infected more quickly if they are surrounded by two infected people rather than just one. Since our estimates are based primarily on transmission events reported during the early stages of outbreaks, we do not explicitly account for such compression and interpret the estimates as *basic* serial intervals at the outset of an epidemic. However, if some of the reported infections occurred amidst growing clusters of cases, then our estimates may reflect effective (compressed) serial intervals that would be expected during a period of epidemic growth. Third, the identity of each infector and the timing of symptom onset were presumably based on individual recollection of past events. If recall accuracy is impeded by time or trauma, cases may be more likely to attribute infection to recent encounters (short serial intervals) over past encounters (longer serial intervals). In contrast, the reported serial intervals may be biased upwards by travel-related delays in transmission from primary cases that were infected in Wuhan or another city before returning home. If their infectious period started while still traveling, then we may be unlikely to observe early transmission events with shorter serial intervals. Indeed, the mean serial interval is slightly higher for the 218 of 301 unique infectors reported to be imported cases.

Given the heterogeneity in type and reliability of these sources, we caution that our findings should be interpreted as working hypotheses regarding the infectiousness of COVID-19 requiring further validation as more data become available. The potential implications for COVID-19 control are mixed. While our lower estimates for *R*_0_ suggest easier containment, the large number of reported asymptomatic transmission events is concerning.

**Figure.**
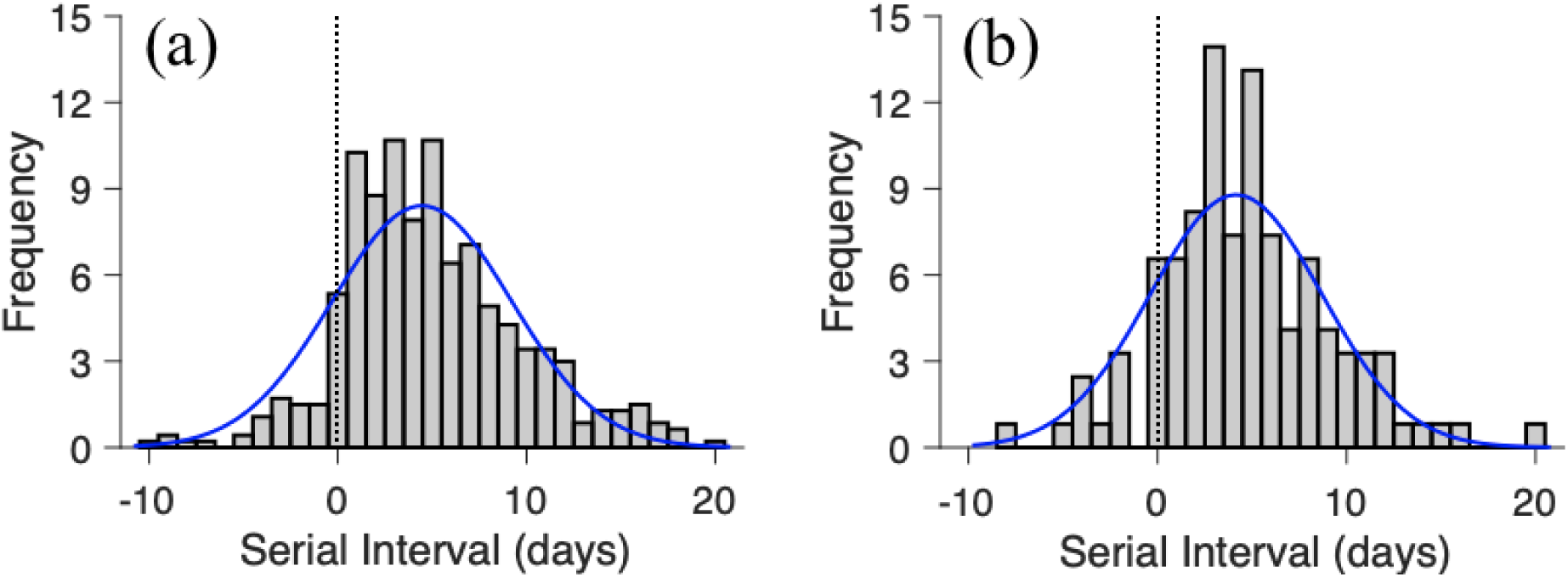
Estimated serial interval distribution for COVID-19 based on 468 reported transmission events in China between January 21, 2020 and February 8, 2020. Bars indicate the number of infection events with specified serial interval and blue lines indicate fitted normal distributions for (a) all infection events (*N* = 468) reported across 93 cities of mainland China by February 8, 2020 and (b) the subset infection events (*N* = 122) in which both the infector and infectee were infected in the reporting city (i.e., the index case was not an importation from another city). Negative serial intervals (left of the vertical dotted lines) suggest the possibility of COVID-2019 transmission from asymptomatic or mildly symptomatic cases.

**Table S1.** Estimated serial interval distributions based on the location of index infection (imported versus local) and the secondary infection (household versus non-household). We assume that the serial intervals follow normal distributions and report the estimated means and standard deviations for all 468 infector-infectee pairs reported from 93 cities in mainland China by February 8, 2020, 122 pairs in which the index case was infected locally, 346 pairs in which the index case was an importation from another city, 104 pairs in which the secondary transmission event occurred within a household, and 180 pairs in which the secondary transmission event was reported as non-household. The rightmost column provides the proportion of infection events in which the secondary case developed symptoms prior to the index case.

| Group | Mean [95 CI%] | SD [95 CI%] | Proportion of serial intervals < 0 |
| --- | --- | --- | --- |
| All (N=468) | 3.96 [3.53, 4.39] | 4.75 [4.46, 5.07] | 12.61% (N = 59) |
| Locally infected index case (N=122) | 3.66 [2.84, 4.47] | 4.54 [4.03, 5.20] | 14.75% (N = 18) |
| Imported index case (N=346) | 4.06 [3.55, 4.57] | 4.82 [4.48, 5.21] | 11.85% (N = 41) |
| Household secondary infection (N=104) | 4.03 [3.12, 4.94] | 4.69 [4.12, 5.43] | 16.35% (N = 17) |
| Non-household secondary infection (N=180) | 4.56 [3.85, 5.27] | 4.80 [4.35, 5.36] | 11.11% (N = 20) |

## Supplementary Appendix

### Data

We collected publicly available data on 6,903 confirmed cases from 271 cities of mainland China, that were available online as of February 8, 2020. The data were extracted in Chinese from the websites of provincial public health departments and translated to English (Table S5). We then filtered the data for clearly indicated transmission events consisting of: (i) a known *infector* and *infectee*, (ii) reported locations of infection for both cases, and (iii) reported dates and locations of symptom onset for both cases. We thereby obtained 468 infector-infectee pairs identified via contact tracing in 93 Chinese cities between January 21, 2020 and February 8, 2020 (Figure S1). The index cases (infectors) for each pair are reported as either importations from the city of Wuhan (*N* = 239), importations from cities other than Wuhan (*N* = 106) or local infections (*N* = 122). The cases included 752 unique individuals, with 98 index cases who infected multiple people and 17 individuals that appear as both infector and infectee. They range in age from 1 to 90 years and include 386 females, 363 males and 3 cases of unreported sex.

## Inference Methods

### Estimating serial interval distribution

For each pair, we calculated the number of days between the reported symptom onset date for the infector and the reported symptom onset date for the infectee. Negative values indicate that the infectee developed symptoms before the infector. We then used the fitdist function in Matlab (19) to fit a normal distribution to all 468 observations. It finds unbiased estimates of the mean and standard deviation, with 95% confidence intervals. We applied the same procedure to estimate the means and standard deviations with the data stratified by whether the index case was imported or infected locally.

### Estimating the basic reproduction number (*R*_0_)

Given an epidemic growth rate *r* and *normally distributed generation times* with mean (*μ*) and standard deviation (*σ*), the basic reproduction number is given by

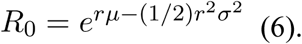

Since we do not know the COVID-19 generation time distribution, we use our estimates of the COVID-19 serial interval distribution as an approximation (Table S1), noting that the serial interval distribution tends to be more variable than the generation time distribution. We assume that the COVID-19 growth rate (*r*) is 0.10 per day [95% CI 0.050-0.16] based on a recent analysis of COVID-19 incidence in Wuhan, China (13). To estimate *R*_0_, we take 100,000 Monte Carlo samples of the growth rate (*r* ∼ *N*(.1,0.028)) and the mean and standard deviation of the serial interval ((*μ,σ*) ∼ *N*(M, Σ) where M = (3.96, 4.75) and 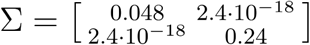). We thereby estimate an *R*_0_ of 1.32 [95% CI 1.16-1.48].

## Model Comparison

We used maximum likelihood fitting and the Akaike information criterion (AIC) to evaluate four candidate models for the COVID-19 serial interval distributions: normal, lognormal, Weibull and gamma. Since our serial interval data includes a substantial number of non-positive values, we fit the four distributions both to truncated data in which all non-positive values are removed and to shifted data in which 12 days are added to each observation (Figure S1 and Tables S2-S3). The lognormal distribution provides the best fit for the truncated data (followed closely by the gamma and Weibull). However, we do not believe there is cause for excluding the non-positive data and would caution against making assessments and projections based on the truncated data. The normal distribution provides the best fit for the full dataset (shifted or not) and thus is the distribution we recommend for future epidemiological assessments and planning.

**Figure S1.**
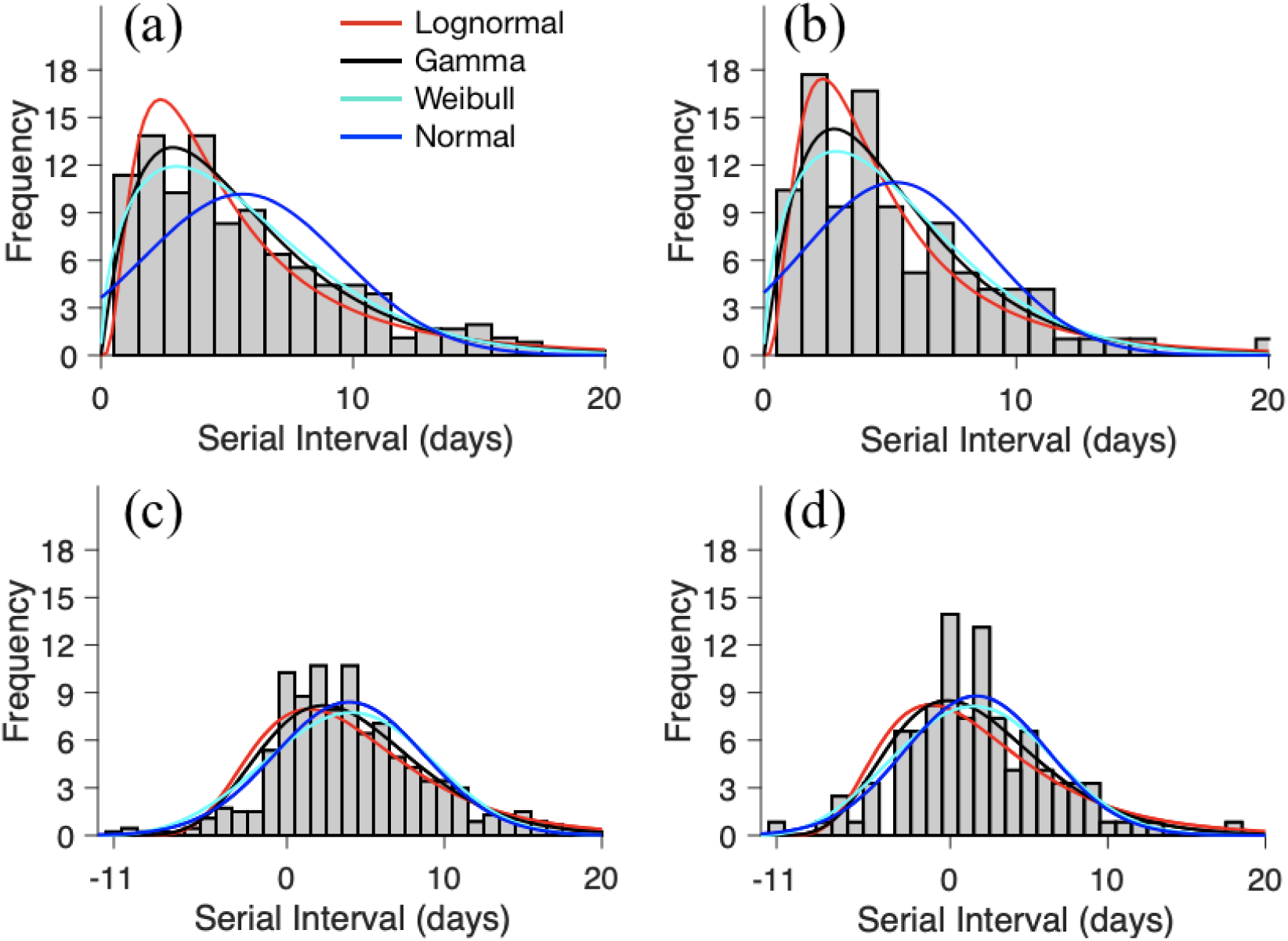
Maximum likelihood distributions fit to transformed COVID-19 serial intervals (468 reported transmission events across 93 cities in Mainland China between January 21, 2020 and February 8, 2020). To evaluate several positive-valued distributions (lognormal, gamma and Weibull), we took two approaches to addressing the negative-valued data. First, we left truncated the data (i.e., removed all non-positive values) for (a) all 468 infection events and (b) the subset of infection events (*N* = 122) in which both the infector and infectee were infected in the reporting city (i.e., the index case was not an importation from another city). Second, we shifted the data by adding 12 days to each reported serial interval for (c) all infection events and (d) the subset of infection events in which both the infector and infectee were locally infected. Bars indicate the number of infection events with the specified serial interval and colored lines indicate the fitted distributions. Parameter estimates and AIC values are provided in Table S4.

**Table S2.** Model comparison for COVID-19 serial intervals based on all 468 reported transmission events in China between January 21, 2020 and February 8, 2020.

| Data | Distribution | Mean/Shape [95% CI] | SD/Scale [95% CI] | AIC |
| --- | --- | --- | --- | --- |
| Original data | Normal (Mean, SD) | 3.96 [3.53, 4.39] | 4.75 [4.46, 5.07] | 2,788.77 |
| Truncated (>0) | Normal (Mean, SD) | 5.62 [5.21, 6.03] | 3.92 [3.66, 4.23] | 2,014.12 |
|  | Lognormal (Shape, Scale) | 2.02 [1.76,2.31] | 2.78 [2.39,3.25] | 1,886.73 |
|  | Gamma (Shape, Scale) | 1.46 [1.38,1.54] | 0.78 [0.72,0.84] | 1,898.63 |
|  | Weibull (Shape, Scale) | 6.25 [5.81,6.72] | 1.50 [1.38,1.62] | 1,892.04 |
| Shifted (+12d) | Normal (Mean, SD) | 15.96 [15.53,16.39] | 4.75 [4.46,5.07] | 2,788.77 |
|  | Lognormal (Shape, Scale) | 9.83 [8.66,11.15] | 1.62 [1.43,1.85] | 2,822.71 |
|  | Gamma(Shape, Scale) | 2.72 [2.69,2.75] | 0.35 [0.33,0.38] | 2,898.24 |
|  | Weibull(Shape, Scale) | 17.66 [17.19,18.14] | 3.56 [3.32,3.80] | 2,806.21 |

**Table S3.**
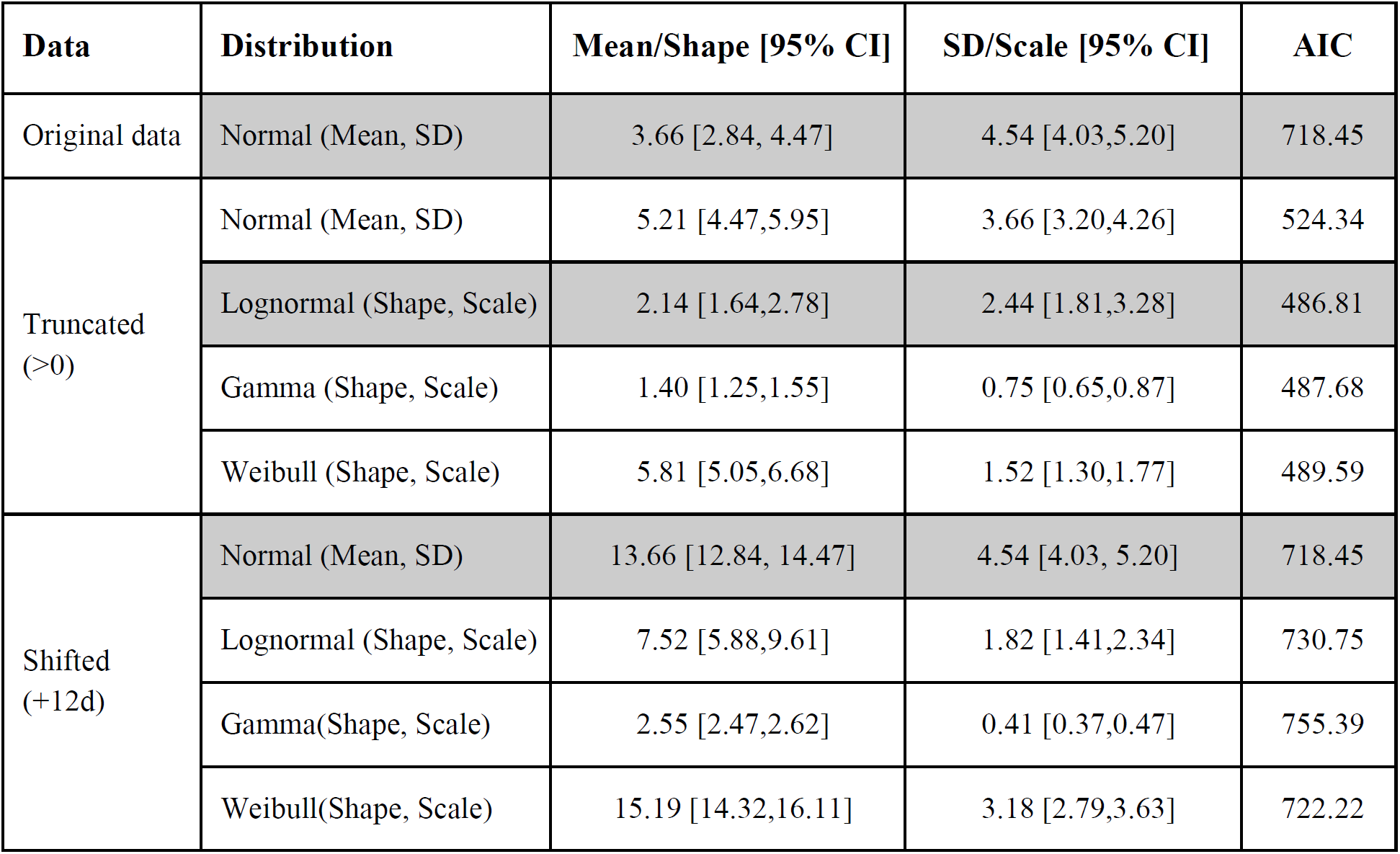
Model comparison for COVID-19 serial intervals based on 122 reported transmission events in China between January 21, 2020 and February 8, 2020 in which both the infector and infectee were infected in the reporting city (i.e., the index case was not an importation from another city).

## Data Availability

All data used in this study is from open access.

## Supplementary Analysis

To facilitate interpretation and future analyses, we summarize key characteristics of the COVID-2019 infection report data set.

### Age distribution

Of the 737 unique cases in the data set, 1.7%, 3.5%, 54.1%, 26.1% and 14.5% were ages 0-4, 5-17, 18-49, 50-64, and over 65 years, respectively. Across all transmission events, approximately one third occurred between adults ages 18 to 49, ∼92% had an adult infector (over 18), and over 99% had an adult infectee (over 18) (Table S4).

### Secondary case distribution

Across the 468 transmission events, there were 301 unique infectors. The mean number of transmission events per infector is 1.55 (Figure S2) with a maximum of 16 secondary infections reported from a 40 year old male in Liaocheng city of Shandong Province.

### Geographic distribution

The 468 transmission events were reported from 93 Chinese cities in 17 Chinese provinces and Tianjin (Figure S3). There are 22 cities with at least five infection events and 71 cities with fewer than five infection events in the sample. The maximum number of reports from a city is 72 for Shenzhen, which reported 339 cumulative cases as of February 8, 2020.

**Table S4.**
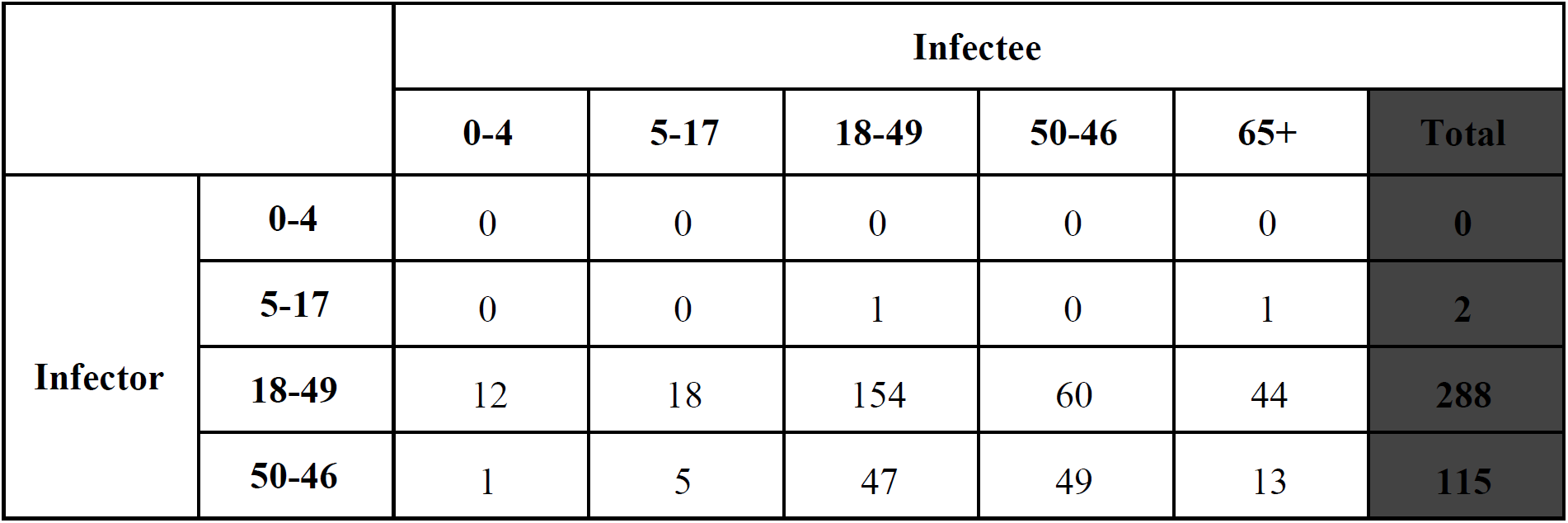
Age distribution for the 457 of 468 infector-infectee pairs. Each value denotes the number of infector-infectee pairs in the specified age combination. Age was not reported for the remaining 11 pairs.

**Table S5.**
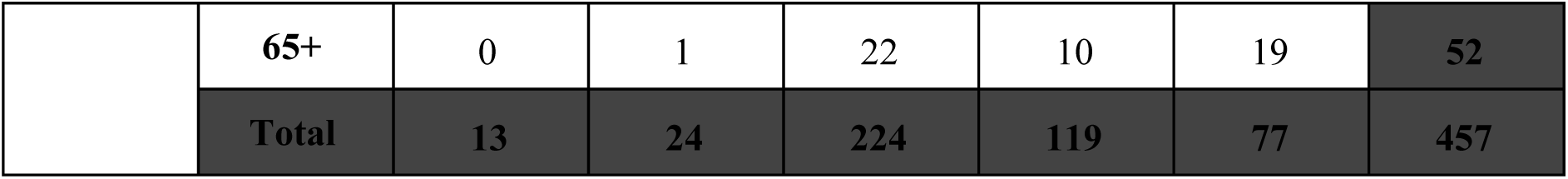
Case report data for 468 COVID-19 infections occurring in 93 Chinese cities by February 8, 2020. Data will be available at https://github.com/MeyersLabUTexas/COVID-19

**Figure S2.**
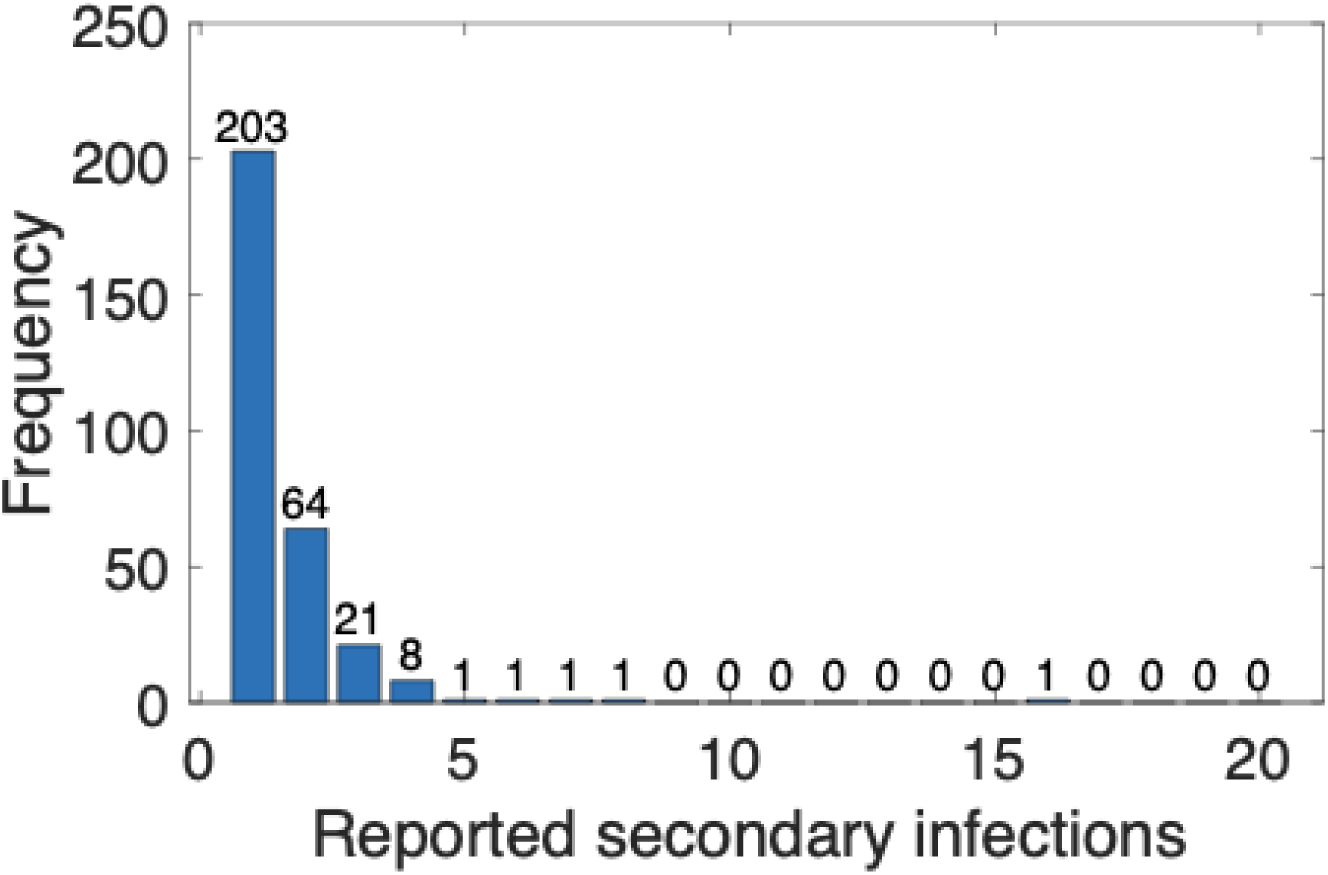
Number of infections per unique index case in the infection report data set. There are 301 unique infectors across the 468 infector-infectee pairs. The number of transmission events reported per infector ranges from 1 to 16, with ∼55% having only one.

**Figure S3.**
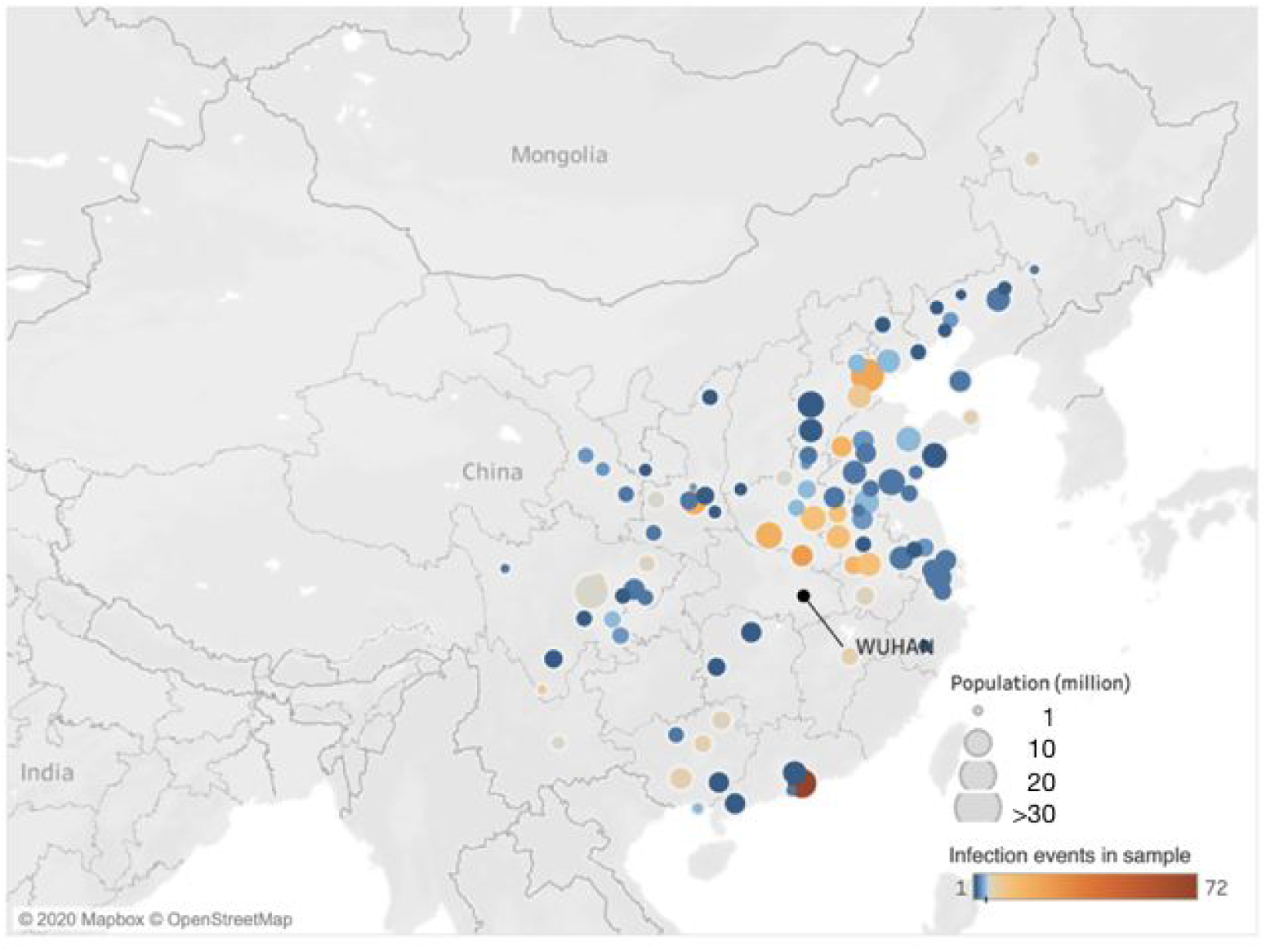
Geographic composition of the infection report data set. The data consist of 468 infector-infectee pairs reported by February 8, 2020 across 93 cities in mainland China. Colors represent the number of reported events per city, which range from 1 to 72, with an average of 5.03 (SD 8.54) infection events. The 71 cities with fewer than five events are colored in blues; the 22 cities with at least five events are colored in shades of orange.

## Acknowledgments

We acknowledge the financial support from NIH (U01 GM087719) and the National Natural Science Foundation of China (61773091).

## Author Bio

Dr. Du is a postdoctoral researcher in the Department of Integrative Biology at the University of Texas at Austin. He develops mathematical models to elucidate the transmission dynamics, surveillance, and control of infectious diseases.

## References

1. WHO | Pneumonia of unknown cause – China. 2020 Jan 30 [cited 2020 Feb 18]; Available from: https://www.who.int/csr/don/05-january-2020-pneumonia-of-unkown-cause-china/en/

2. Organization WH, Others. Coronavirus disease 2019 (COVID-19): situation report, 30. 2020; Available from: https://www.who.int/docs/default-source/coronaviruse/situation-reports/20200219-sitrep-30-covid-19.pdf?sfvrsn=6e50645_2

3. Cowling BJ, Leung GM. Epidemiological research priorities for public health control of the ongoing global novel coronavirus (2019-nCoV) outbreak. Euro Surveill [Internet]. 2020 Feb 13; Available from:http://dx.doi.org/10.2807/1560-7917.ES.2020.25.6.2000110

4. Giesecke J. Modern infectious disease epidemiology. CRC Press; 2017.

5. Svensson A. A note on generation times in epidemic models. Math Biosci. 2007 Jul;208(1):300–11.

6. Wallinga J, Lipsitch M. How generation intervals shape the relationship between growth rates and reproductive numbers. Proc Biol Sci. 2007 Feb 22;274(1609):599–604.

7. Vink MA, Bootsma MCJ, Wallinga J. Serial Intervals of Respiratory Infectious Diseases: A Systematic Review and Analysis [Internet]. Vol. 180, American Journal of Epidemiology. 2014. p. 865–75. Available from: http://dx.doi.org/10.1093/aje/kwu209

8. Kuk AYC, Ma S. The estimation of SARS incubation distribution from serial interval data using a convolution likelihood. Stat Med. 2005 Aug 30;24(16):2525–37.

9. Lipsitch M, Cohen T, Cooper B, Robins JM, Ma S, James L, et al. Transmission dynamics and control of severe acute respiratory syndrome. Science. 2003 Jun 20;300(5627):1966–70.

10. Cowling BJ, Park M, Fang VJ, Wu P, Leung GM, Wu JT. Preliminary epidemiological assessment of MERS-CoV outbreak in South Korea, May to June 2015 [Internet]. Vol. 20, Eurosurveillance. 2015. Available from: http://dx.doi.org/10.2807/1560-7917.es2015.20.25.21163

11. Park SH, Kim Y-S, Jung Y, Choi SY, Cho N-H, Jeong HW, et al. Outbreaks of Middle East Respiratory Syndrome in Two Hospitals Initiated by a Single Patient in Daejeon, South Korea. Infect Chemother. 2016 Jun;48(2):99–107.

12. Jung S-M, Akhmetzhanov AR, Hayashi K, Linton NM, Yang Y, Yuan B, et al. Real time estimation of the risk of death from novel coronavirus (2019-nCoV) infection: Inference using exported cases [Internet]. Available from: http://dx.doi.org/10.1101/2020.01.29.20019547

13. Li Q, Guan X, Wu P, Wang X, Zhou L, Tong Y, et al. Early Transmission Dynamics in Wuhan, China, of Novel Coronavirus-Infected Pneumonia. N Engl J Med [Internet]. 2020 Jan 29; Available from: http://dx.doi.org/10.1056/NEJMoa2001316

14. Tuite AR, Fisman DN. Reporting, Epidemic Growth, and Reproduction Numbers for the 2019 Novel Coronavirus (2019-nCoV) Epidemic [Internet]. Annals of Internal Medicine. 2020. Available from: http://dx.doi.org/10.7326/m20-0358

15. Wu JT, Leung K, Leung GM. Nowcasting and forecasting the potential domestic and international spread of the 2019-nCoV outbreak originating in Wuhan, China: a modelling study. Lancet [Internet]. 2020 Jan 31; Available from: http://dx.doi.org/10.1016/S0140-6736(20)30260-9

16. Zhao S, Gao D, Zhuang Z, Chong M, Cai Y, Ran J, et al. Estimating the serial interval of the novel coronavirus disease (COVID-19): A statistical analysis using the public data in Hong Kong from January 16 to February 15, 2020. medRxiv [Internet]. 2020; Available from: https://www.medrxiv.org/content/10.1101/2020.02.21.20026559v1.abstract

17. Nishiura H, Linton NM, Akhmetzhanov AR. Serial interval of novel coronavirus (2019-nCoV) infections [Internet]. Infectious Diseases (except HIV/AIDS). medRxiv; 2020. Available from: https://www.medrxiv.org/content/10.1101/2020.02.03.20019497v1.abstract

18. Kenah E, Lipsitch M, Robins JM. Generation interval contraction and epidemic data analysis. Math Biosci. 2008 May;213(1):71–9.

19. Fit probability distribution object to data - MATLAB fitdist [Internet]. [cited 2020 Feb 19]. Available from: https://www.mathworks.com/help/stats/fitdist.html

